# A qualitative investigation of young people’s experiences and views of Early Support Hubs

**DOI:** 10.1101/2025.09.05.25335175

**Authors:** Lily Wright, Jessica L. Griffiths, Rebecca Appleton, Sadiya Begum, Connor Clarke, Nima Cas Hunt, Hannah Kate Lewis, Phoebe Barnett, Anam Bhutta, Eva Driskell, Julian Edbrooke-Childs, Andrew Grundy, Isabel Hanson, Emma Maynard, Lizzie Mitchell, Rob Saunders, Polly Waite, Brynmor Lloyd-Evans, Kylee Trevillion, Sonia Johnson

## Abstract

**Background:** Youth is a period of elevated risk for mental ill health, yet young people often do not receive timely support. Barriers can include high clinical thresholds for treatment and long waiting lists, as overstretched statutory services can struggle to meet high demand. The Early Support Hubs available in some parts of England are a potentially promising model to increase access to support. These are community-based services offering open-access, holistic support for 11–25-year-olds without a referral. However, there is no standardised model and considerable variation in the support offered, highlighting the need for research to explore how Early Support Hubs operate, whether they are meeting the mental health and wellbeing needs of young people, and potential areas for improvement.

**Aims:** To explore young people’s experiences of using Early Support Hubs for mental health or wellbeing support, and their views on best practice within these services.

**Methods:** We conducted semi-structured interviews with 20 young people aged 16-25 years, who had used Early Support Hub services across England. Data were analysed using codebook thematic analysis.

**Results:** Aspects of hubs that were valued by young people included: easy accessibility; holistic approaches which go beyond clinical interventions; a sense of community, friendship and consistency; and youth-led philosophies. Limitations of the hub model included them being little known in local areas, lack of capacity to address more acute and complex mental health needs, and the limited scale of the services.

**Conclusion:** Early Support Hubs appear to be valued by young people and have potential to be an adjunct to clinical services to help increase access to mental health support for young people. Evidence on populations served, what support they receive, and outcomes following support are needed to assess whether there is a policy case for wider roll out.

## Introduction

### Children and young people’s mental health and help-seeking

Youth, including adolescence from ages 10-19 years (1) and young adulthood from ages 19-25 years (2), is a period of high risk for mental health problems (3). Over one-third of all lifetime cases of mental disorders emerge by age 14, nearly half (48.4%) by age 18, and nearly two-thirds (62.5%) by age 25 (4). In England, over a quarter of young adults (aged 16 to 24) were identified with a common mental health condition (APMS, 2025), and overall rates of young people experiencing mental health difficulties have increased over the past two decades (5).

Whilst some mental health problems do resolve, many persist into later adulthood (6). Moreover, mental health difficulties in childhood and adolescence predict negative outcomes, including lower levels of life satisfaction and health-related quality of life, alongside lower levels of academic success and future family difficulties (i.e. marriage stability and family income) (7,8). This indicates a considerable cost burden for both individuals and society (6,9).

Young people face various barriers both to seeking professional help, and to accessing appropriate support once they do. Evidence from both UK and international studies shows these barriers include stigma and embarrassment, confidentiality concerns, limited knowledge of how to identify problems and seek help, and negative beliefs about the relevance, effectiveness and consequences of professional support (10–13). In the UK, structural barriers include increasing thresholds for access to treatment and long waiting lists, as overstretched statutory services attempt to meet growing demand (14,15), as well as a lack of available or appropriate services (e.g., in rural areas or for those needing specialist support). These may also discourage professional help-seeking and delay or prevent access for those who do seek support (16,17).

UK and international evidence indicates that certain demographic groups may face greater barriers to accessing mental health care, including minoritised ethnic groups, younger children, boys, and young people identifying as lesbian, gay, bisexual, transgender, queer and questioning (LGBTQ+) (12,18–24). Child and Adolescent Mental Health Services (CAMHS) in England, funded publicly through the National Health Service (NHS), received over 1.2 million new referrals in 2022 alone, showing a 53% increase from 2019 (25), and one in five children are waiting over six months for contact with a mental health specialist (5). This exceeds care quality guidelines, which recommend that all non-urgent referrals begin treatment within four weeks, with shorter timeframes for young people experiencing psychosis, needing community-based eating disorder treatment, or presenting to crisis services (26).

### The role of Early Support Hubs

Early Support Hubs are a potential service model to increase access to mental health and wellbeing support for young people (6). Informed by the Youth Access model (27), established in 1998, hubs seek to provide open access support for 11–25-year-olds, predominantly on a drop-in basis, and accept self-referrals without the need for a referral from a professional (28–30).

Recognising that mental health does not exist in a vacuum, the Youth Access model also moves away from a diagnostic focus, instead adopting a whole life approach to wellbeing and considering emotional, social and economic contributors (30). Therefore, in addition to drop-in mental health support and therapy offered at some hubs, they may also provide advice and support around wider wellbeing issues which affect mental health, such as substance use, sexual health, employment, housing and financial circumstances (28). This aligns with calls from NHS England’s ‘Future in Mind’ report to expand ‘one-stop-shop’ wellbeing services – models which offer a range of different types of support in a single location (31). Similar open-access early intervention youth models have been established internationally, such as ‘Jigsaw’ in Ireland (32), ‘Headspace’ in Australia (33,34), and @ease in The Netherlands (35), although these generally have a more clinical focus than Early Support Hubs.

In the UK context, the Early Support Hub model reflects calls from the ‘Our Minds Our Future’ manifesto, developed by young people across England, to reduce long waiting lists and avoid arbitrary symptom thresholds which limit access to services (30). While expanding early mental health support in schools has been a priority in recent years (36), community-based Early Support Hubs may help to address barriers to school-based services, such as Mental Health Support Teams, by reaching young people in alternative education. Early Support Hubs also align with the NHS 10 Year Health Plan which seeks to expand community-based mental health services and create comprehensive offerings reaching across adolescence and young adulthood (37). This could address difficulties faced when reaching the upper age limit of CAMHS, for example in adapting to the individualised, diagnostic-based approach of adult services following the more family-oriented and holistic approach of CAMHS (38), or their current care finishing without a transition to adult services, despite ongoing clinical need (39,40).

Around 70 Early Support Hubs operate across England, and funding has recently been made available by the Department of Health and Social Care to expand provision at 24 existing hubs (41). However, a greater understanding of what they are aiming to achieve and what support they deliver is needed in order to be able to evaluate them and develop a model of best practice. Not all hubs subscribe to the Youth Access model, and hubs are commissioned from a variety of local authority and voluntary sector providers (28). Therefore, the population served, and types of support being provided are unlikely to be consistent, and the extent to which they address the mental health needs of a variety of young people is uncertain.

One previous qualitative study explored the experiences of staff, young people, and parents/carers at an Early Support Hub (42). It identified the importance of accessibility and flexibility, a non-clinical model and environment, approachable staff, partnership working, and promotion of positive mental health and wellbeing (42). However, they recruited participants from a single hub in a disadvantaged area in North West England, and given the likely variation in support delivery across different hubs, it is unclear if these findings would apply to other contexts.

### The current study

In this study, we aimed to explore the perspectives of young people who have accessed Early Support Hubs in England. We hope this will provide evidence to inform policy around delivering early interventions in community settings. The study objectives were:

1. To explore the experiences of young people who have used Early Support Hubs for mental health or wellbeing support.
2. To explore young people’s views on what is helpful about Early Support Hubs and what makes for a good service in this context.

## Methods

This study was conducted by the NIHR Policy Research Unit in Mental Health (MHPRU) (43), by researchers based at University College London (UCL) and King’s College London (KCL). The MHPRU is funded via the National Institute for Health and Care Research to conduct research to meet policymaker need.

This project was led by an MSc student (LW), supervised by RA, KT, and SJ, and supported by a working group comprising academic and lived experience researchers, and clinicians. The working group contributed to a broader programme of research on Early Support Hubs, led by the MHPRU, which included literature reviews (44), a quantitative evaluation of Early Support Hubs, and qualitative studies exploring the experiences of hub managers, frontline staff, and young people. The full working group met regularly to discuss the overall programme of research, while a smaller subset held weekly meetings to focus specifically on the qualitative components, including this study.

### Lived experience involvement

The working group included six lived experience researchers: four young people with lived experience of using mental health services, a parent with lived experience of supporting their children with mental health difficulties (ED), and a MHPRU lived experience research fellow (HKL). The ways in which they were involved were attending working group meetings throughout the project and contributing to all stages of the study, including design, data collection, analysis, and write-up.

In recognition of criticisms (45) of the COREQ guidelines for qualitative research (46), we used the COREQ checklist as a prompt for detailed and transparent reporting rather than applying it prescriptively.

### Ethical considerations

Ethical approval was received from University College London Research Ethics Committee: 27031/001. Although hubs provide services for young people aged 11-25, all participants in this study were aged 16-25 and therefore able to provide their own informed consent to participate. Whilst interviews were not intended to elicit detailed accounts of individual mental health or difficult circumstances, a protocol was developed to manage any potential distress. Participants could pause or stop the interview at any point, and the interviewer asked for feedback on their experience of the interview and signposted to relevant organisations (e.g. mental health charities).

### Study design

Individual semi-structured interviews were conducted. The study adopted a critical realist perspective, assuming participants’ comments provide access to an underlying reality of experiences, motivations and meaning, but these are not independent of broader sociocultural processes, including those at play within our research processes (47).

### Setting

We recruited from organisations linked with the 24 Early Support Hubs receiving a share of the Department of Health and Social Care funding (41).

### Eligibility

Young people were eligible if they were aged 16-25 years, had capacity to consent, and reported having used, or attempted to use, an Early Support Hub within the last two years for any type of mental health, wellbeing or stress support.

### Recruitment

To ensure a diverse sample, we aimed to recruit around 20 participants. Conducting data collection and analysis in parallel, we planned to stop recruitment once data was sufficiently rich and complex to address the study objectives (48). Maximum variation sampling (49) was used to ensure diversity in age, ethnicity, gender, sexual orientation, and education/employment status. This was important since hubs may play an important role in engaging marginalised groups who are less likely to access statutory or school-based services, such as LGBTQ+ individuals or those from minoritised ethnic backgrounds (50,51).

Recruitment materials were circulated via the hubs, containing researcher contact details for participants to express interest. If given permission by the young person, staff also shared service user contact details with the research team for us to contact directly. Members of the research team (LW, RA, HKL, CC, JG) also made site visits, meeting with staff to encourage recruitment. Participants were offered £30 shopping e-vouchers to thank them for their time.

### Topic guides

Semi-structured interview topic guides were collaboratively developed by the research team. Questions included: reasons for hub use; timeframe of hub use; experiences of accessing and using hubs; and views on what makes an ideal Early Support Hub (see Appendix A for full topic guide). Interviews were designed to last approximately 45-60 minutes.

### Data collection

Most interviews (n = 11) were conducted by LW, a female White British MSc student, while the remaining interviews (n = 9) were conducted by other members of the research team, including one female White British lived experience research fellow (HKL), a female White British senior lecturer and deputy director of the MHPRU (KT), two female White British postdoctoral researchers (RA, JG) and a male White British research assistant (CC).

Before each interview, the researcher assessed the young person’s capacity to give informed consent by reviewing the participant information sheet, checking participants’ understanding, and providing the opportunity to ask questions. Depending on preference, written or audio-recorded verbal consent was obtained. Participants also completed an online demographics survey, collecting optional data on age, education and employment status, ethnicity, gender, and sexual orientation.

Interviews were audio-recorded and transcribed directly using Microsoft Teams, whether they were conducted online or in-person. Transcripts were reviewed and amended by a researcher (LW, JG, CW, HKL) to ensure accuracy and remove identifying information. There were some instances of unclear audio, though these were infrequent and did not meaningfully affect the overall quality or completeness of the transcripts.

### Data analysis

Transcribed data were analysed using NVivo version 14 for Windows software. Guided by principles of Braun and Clarke’s thematic analysis (52,53), we adopted a codebook approach whereby themes identified in the data were generated and charted into a coding frame. Themes were generated using both an inductive (i.e. new themes observed directly from the text) and deductive (i.e. themes derived from policy-relevant topics of interest) approach. The coding frame was used to record and chart the developing analysis, as well as guiding data coding for the multiple coders (54). The key steps of our analytical approach are outlined below:

- Initial data familiarisation involved detailed reading and review of the transcripts and interview recordings.
- Independent coding was then undertaken for two transcripts by two coders (LW, KT), and an initial coding frame was developed based on consensus discussions in relation to the identified themes in the data.
- KT, AG and RA delivered training on how to code qualitative data using a thematic synthesis approach to four lived experience researchers in the working group. The lived experience researchers then all independently coded one of the research interviews before meeting again with KT to discuss and compare their coding; this resulted in further review and updating of the initial coding frame.
- The coding frame was then applied to the remaining data, and new themes iteratively added, refined and combined throughout the coding process (LW, JG, HKL). At several stages in the coding process the developing coding frame was reviewed to check its suitability and to facilitate further iterations.
- The final coding framework was used to write up each theme, including illustrative quotes, to write an analytical narrative around the data (LW, JG). A meeting was held between three of the academic researchers (RA, KT, JG) and four lived experience researchers to discuss the interpretation of the final coding frame and write-up of the themes. The write-up was revised accordingly, and further feedback was sought from the wider working group on the draft of the paper.

## Results

### Participants

Twenty young people were recruited from eight Early Support Hubs across England. Each of these hubs offered drop-in advice services, and most also offered more structured courses of therapy or counselling, either on an individual or group basis. Interviews were conducted between June and November 2024 and ranged from 11-72 minutes in length. Full demographic details of participants are presented in Table 1.

**Table 1.** Participant demographics.

|  |  |
| --- | --- |
| **Age (years), mean (SD)** | 19.9 (2.7), range 16-25 |
| **Gender, N** |  |
| Male | 10 |
| Female | 8 |
| Non-binary | 1 |
| Other | 1 |
| **Ethnicity, N** |  |
| White British | 15 |
| Any Black background | 4 |
| Any Asian background | 1 |
| **Sexual orientation, N** |  |
| Heterosexual/straight | 7 |
| Bisexual | 5 |
| Other | 4 |
| Gay/lesbian | 3 |
| Prefer not to say | 1 |
| **Employment status, N** |  |
| Employed part-time | 6 |
| Not in education, employment, or training | 5 |
| School/sixth form/college student only | 4 |
| School/sixth firm/college student and part-time employed | 3 |
| Employed full-time | 1 |
| University student and part-time employed | 1 |
| **Length of engagement with hub(s), N** | Range: 6 weeks – 9 years |
| <1 year | 4 |
| 1 to <2 years | 5 |
| 2 to <3 years | 1 |
| 3 to <4 years | 1 |
| 4 to <5 years | 4 |
| 6+ years | 3 |
| Not reported/unclear | 2 |

Interview data showed most participants (n = 18) reported mental health difficulties, though these were not always the main reason for accessing hubs, with some first seeking help due to social or economic issues such as housing or benefits problems. Anxiety and depression/low mood were the most commonly experienced symptoms, but other perceived or diagnosed difficulties included obsessive compulsive disorder, eating disorders, sleep problems, anger management difficulties, trauma-related difficulties, and substance use issues. Over a third (35%) of the sample reported that they were autistic, although it was uncertain how many had received a formal diagnosis. Participants reported engaging with hub support for durations ranging from 6 weeks up to 9 years, with the majority (70%) reporting receiving hub support for over a year.

### Overview

We identified five themes capturing participants’ experiences of and views on Early Support Hubs: i) ‘beyond clinical interventions: a holistic approach to wellbeing’; ii) ‘accessibility’; iii) ‘friendship, community, and consistency’’; iv) ‘youth-centred approach’; and v) ‘hubs as limited by their scale and reach’. Each theme is described below, accompanied by illustrative quotes.

### Theme 1: Beyond clinical interventions: a holistic approach to wellbeing

A key aspect of the Early Support Hub model which participants valued was the holistic and non-clinical approach which extended beyond traditional mental health interventions.

Participants sought support from hubs for various difficulties, including mental health problems, substance use, domestic violence, young parenthood, sexual health, LGBTQ+ identities, and financial, legal, education, employment, and housing difficulties. Some participants reported that hubs offered drop-in support, and some described access to more structured time-limited individual or group interventions at hubs, such as therapy; counselling; information, advice and guidance; and physical activity programmes. The maximum number of therapy/counselling sessions reported by participants mostly ranged between 6-14. In this context, participants highlighted the benefit of the ‘one-stop-shop’ model, where they could access several types of support in one place:

> *“It’s so convenient. It’s like popping into Asda, needing food and cleaning products and dog food.”* (YP7)

Crucially, hubs were also well connected with other services, allowing them to signpost to support outside their remit:

> *“[Hub staff] also get in touch with services they think can help, and they’ll get in touch with GPs, things like that, if they need to. Especially if you feel unable to do it yourself… they redirect you to services that they think will help.”* (YP3)

In line with this, participants did not view their mental health as an isolated entity which could only be improved through formal psychological therapies. Many instead emphasised the benefit of more holistic support to improve mental health and general wellbeing:

> *“Not everyone wants to get a psychologist. It’s not about that. Sometimes it’s just about popping in and knowing that there’s someone to talk to.”* (YP2)

Participants valued being able to choose what hub support to engage with, based on their individual needs, rather than this being determined by a diagnosis or limited to a single treatment pathway offered by professionals:

> *“Not saying [diagnosis] doesn’t help, but there’s too many labels of sort of everything of this kind in this world today, and I just see myself as a guy… so you go there just to be seen as, ‘oh, it’s just [name], it’s this person’.”* (YP2)

Young people also liked the non-clinical and homely environment of hubs. They noted how buildings resembling houses, colourful decor, and comfortable furniture contributed to feelings of safety, contrasting from experiences of discomfort in more clinical spaces such as General Practitioners (GPs) or adult mental health services:

> *“When you go to a GP clinic, no matter how nice they are to you, it feels like as a clinic, it doesn’t feel like it’s, you know, it doesn’t feel homely, you don’t feel 100% safe, in my hub like it’s like the way that the furniture is laid out… it feels a lot more homely.”* (YP9)

Whilst the comprehensive and non-clinical model worked well for most, it was perceived as too general to address more complex mental health difficulties such as post-traumatic stress disorder or obsessive-compulsive disorder. Some participants presented this as a critique of the hubs, wanting there to be more specialist staff *“rather than having everyone know a little bit about a lot, but not a lot about a little”* (YP11). Conversely, another participant felt positively about receiving more formal interventions elsewhere, since this allowed the hub to remain a *“happy safe place separate from working through my trauma”* (YP7).

### Theme 2: Accessibility

Experiences of accessing Early Support Hubs were two-fold. Initially, young people faced barriers to identifying hubs as useful sources of support, but thereafter generally described easy accessibility as key to their success.

Hubs were not perceived as well known in their local communities, and several young people described feeling anxious when first visiting due to a lack of prior knowledge and expectations, and difficulties identifying hub locations due to unclear signage and the residential appearance of some hubs:

> *“It feels a bit closed off when it’s just like it’s not a see-through door… it’s good with like with glass doors because then you can kind of like feel like you’re you can see what’s going on and you don’t feel like, oh, what do I do?”* (YP4)

A couple of participants described hub advertisements as focusing on drug and alcohol or sexual health support. In these instances, participants with different support needs perceived the hubs as either not being relevant to their needs or potentially stigmatising, which could be potential barriers to accessing hub support:

> *“They were basically where you would go to get like sanitary towels or condoms or tampons, things like that, and not as a thing that you’d go to if you were down or depressed…that’s kind of what stopped me from using them to begin with…I was a bit like, I don’t want to be seen as the person that goes there.”* (YP8)

However, for those who chose to still access support from hubs, this anxiety generally eased over time due to the hubs’ welcoming, inclusive atmosphere and their flexible, drop-in nature – allowing young people to engage on their own terms without needing to commit to a fixed programme:

> *“It actually took me a while to actually make the first step to come to one of the sessions. But the first session I came to was the Wednesday one. I decided to just give it a go and if it wasn’t for me then I could always back out. So it was quite nerve wracking at first, but as soon as I got there, everyone was very welcoming. It was other people’s first time as well, so I got to know people.”* (YP12)

Hubs were generally seen as physically accessible, with many located in town centres, close to schools and colleges, and with good public transport links. Some services also had hubs in multiple locations to increase accessibility, and several were described as wheelchair accessible.

> *“I’m quite lucky in there is a bus I can get that drops me off a few minute walk from it or it is, I think it’s something like 30 minute walk from my house to get there… and obviously I was going into college, which meant I could walk from college to there within 5 minutes. It was really simple to get from appointments… it made it so much easier to have it fairly nearby.”* (YP11)

However, one participant described facing difficulties accessing their hub due to gang-related violence in the local area, including at the hub’s location, which at times required police intervention:

> *“I had a few difficulties, especially when erm there was a lot of gang wars… I may or may not have been jumped around here a few times, even outside the youth club… they would try and come here, cause trouble and then the police will have to shut them down.”* (YP19)

Young people valued being able to access drop-ins without a need for a referral, the timely and flexible nature of the support, the lack of focus on clinical thresholds, and having the option to engage with support face-to-face or online. Hub access was often through online self-referrals, which participants generally agreed *“makes it ten times easier to access”* (YP11).

> *“We can like self-refer and then you can also join a group… if you refer and you’re like I need something now… it just makes it a bit more accessible and I think that’s the best kind of part about it… it’s not just put yourself on a waiting list and then we’ll talk to you in six months.”* (YP6)

One participant described how language barriers made it challenging for them to engage with hub support, and how they found it helpful to have others present who spoke the same language. Another participant noted that young people could access translators at hubs when needed.

> *“They do occasionally have access to translators. English is my first language, so again that doesn’t apply to me, but there have been people that needed it, and they have had access.”* (YP7)

Another access barrier related to parental consent requirements for younger people. One participant stated that younger children had to be signed into the hub by their parents, but expressed a wish that *“it would be like a proper open access youth club where they don’t have to sign you in”* (YP20). Another 16-year-old participant shared that they were able to access hub support without parental consent, and that in some cases young people could engage in counselling at the hub without parental consent. They suggested that the age requirement for accessing support without needing parental consent should be lowered, as it can be difficult for some young people to discuss mental health challenges with their families:

> *“I do feel like people need to lower the thing saying you need to get your parents’ permission because sometimes the parents don’t know about the kids’ issues because they were brought up on the fact of ’you shouldn’t talk about them’.”* (YP16)

### Theme 3: Friendship, community and consistency

Many young people valued the community and friendships provided by hubs, making them an enjoyable place to be. This theme emerged in relation to dynamics with other young people, staff members, and the hub model itself.

Hubs were social spaces for many, providing unique opportunities to form friendships with like-minded individuals of a similar age, with a shared identity, or who had experienced similar difficulties. Some young people described attending groups or just ‘popping in’ to enjoy the hub community, as opposed to accessing the hub to seek help for a specific difficulty:

> *“…if I’m not doing anything on a day during the week, if I’ve literally got nothing to do, I’ll most likely go [to the hub] and a couple of other friends will probably be knocking around or I’ll just go there to actually help make cups of teas and things for people.”* (YP1)

The consistent provision of support by hubs meant that some participants viewed hubs as a longer-term support system which could be used flexibly and reliably over several years, as opposed to a service providing only short-term interventions:

> *“When I left they still gave me someone I could reach out to and contact… they’ll still reach out and contact me, anything that I might be interested in, and if I needed to arrange more appointments, I could go to them… you still have that foot in the door, in case you do need to go back.”* (YP11)

A sense of community and wanting to give back to hubs was also reflected in some young people’s comments around volunteering at their hub or wanting to work for them in the future:

> *“They’ve actually maybe encouraged me and taught me that maybe I would like to deliver the same sort of help to people like myself once I’ve got myself sorted. They do offer opportunities like that.”* (YP2)

Young people described developing strong rapport with staff, attributing this to their informal, less clinical approaches, approachable and relatable demeanours, and genuine efforts to get to know hub users as individuals. This contrasted from more formal and structured, clinically focused encounters experienced at other services:

> ***“****In more professional systems you can feel quite like a case number… [at hub] you generally get to know the staff individually as people… so you make connections and you genuinely feel like a person, you feel like you’re actually talking to a person, you feel valued, you feel cared about.”* (YP2)

This warm, welcoming approach extended to one participant’s young child, who they occasionally brought to the hub with them. The participant felt that over time their child formed a strong bond with staff, and that they became *“like another family to him”* (YP7).

Some young people valued that staff had lived experiences similar to their own, such as growing up in the local area or having shared identities. They felt his helped them to connect with staff and feel understood:

> *“And the fact that they’ve all been hand selected for having like the same disabilities as us and the same sexualities as us, but just grown up, it really puts you in a safe environment where you know that they won’t judge you either because they’re just in the same shoes as you, just like everyone else in the places.”* (YP12)

However, in the context of such a tight-knit community where young people could usually talk to familiar staff members in regular appointments, drop-ins, and groups, some participants did recall being upset by sudden and sometimes frequent staffing changes. This disrupted continuity of care and led to session time being used to rebuild trust:

> *“There have been times where staff I really like and got on with well had to leave and that was quite upsetting for me, ’cause again, like, I built those relationships and it’s hard for me to trust staff at mental health services… it can be challenging for me to then open up to a new staff member and to have to kind of say things all over again.”* (YP5)

### Theme 4: Youth-centred approaches

Young people emphasised the importance of hubs having a youth-centred ethos, particularly in terms of choice around support, dynamics with staff, and involvement with decision-making about hub design and delivery at large.

Hubs encouraged young people to take charge of their wellbeing journeys from the first point of contact by accepting self-referrals and allowing choice over the level of family involvement, with preferences regarding family involvement varying between participants:

> *“Doctors can refer you but they prefer if you refer yourself… if you refer yourself, then you want the help kind of thing.”* (YP8)
>
> *“I made it very, very clear that I wanted to be the first point of contact…my dad came to the interview for [hub], not by choice. I would have rather done it alone…and then my dad left and I was there alone and I essentially re-explained a lot of the situations… [staff member] believes what I’m saying, which is quite crucial for me because if I have no purpose to lie, it really irritates me when I do get accused of it, so it’s nice to know that he does trust me.”* (YP11)

One participant described there being occasions when hub staff contacted their mother about their behaviour, such as when they became *“a little bit too crazy… too upset”*. While this could feel *“overwhelming*”, as it could lead to them *“getting into trouble”*, the young person also stated *“if she has to know, she has to know”* (YP20).

Most participants were given choice around the type of support they accessed and were not pressured to agree with professional opinions or pursue particular interventions. Instead, support was affirming, which was perceived as fundamental within youth contexts where often service users are not treated as equal-decision makers.

> *“A lot of youth places do just kind of have that, like, ‘I’m in charge, you’re the young person’, but this is kind of like we’re all in charge here.”* (YP6)

However, choice of support was not universally described by all participants:

> *“With my autism, I can find it difficult to kind of understand things and to change my thoughts… I kinda wish that I was offered a choice of what therapy I wanted. I was kind of just given CBT.” (YP5)*

Beyond decisions about individual support, young people felt valued by the fact that hubs actively reached out to them to help shape the service, for example through youth participation groups which guided decisions about building designs, support offerings and recruiting new staff members:

> *“… we all got a say in what that looked like, like they asked us what we wanted in there and we kind of we designed that room.”* (YP5)
>
> *“Young people will get to do an interview [with potential employees] where we ask like just some questions and we get to have a vibe of people, which I think is so important because obviously these are the people who are going to be helping young people.”* (YP6)

Some participants also described taking on ‘Young Ambassador’ or ‘Young Commissioner’ roles at the hubs, where they participated in projects involving co-production, advocacy, and outreach to other organisations.

> *“We do a lot of stuff on youth voice and co-production as well as the mental health side of things.”* (YP15)

The sense of autonomy fostered was also reflected in young people’s perceptions that hubs were often not the sole reason for improvements in their wellbeing or circumstances, but rather hubs supported young people to help themselves:

> *“They’ve always been quite supportive in [you] doing things for yourself… they’ve kind of helped me in that way discover like myself and what I can do to make myself feel better and be better and be healthier.”* (YP8)

### Theme 5: Hubs as limited by their scale and reach

With a consensus that the hub model generally worked well, a desire to extend the scale of hubs emerged as a prominent theme, particularly regarding their size and number, the time-limited nature of support, and the age restrictions for access.

Some participants described how, due to the small size of hub buildings, drop-ins were busy, sometimes cramped, and could be noisy, which at times felt overwhelming. Additionally, there were not always separate rooms available for more private conversations:

> *“If I’m wanting to speak about something particularly sensitive, I may not necessarily want to do that when it’s so busy because like, there are people that could literally be sat a few feet away from you and they will hear you.”* (YP7)

Busy drop-ins sometimes led to long wait times. Whilst most were seen within the hour, one participant recalled being unable to receive same day support on several occasions. Another young person felt that even a short wait could lead to someone deciding to leave the hub:

> *“When you’re in that position, you kind of have a million thoughts going through your head at once, so even just like five minutes is enough to make someone walk out the door.”* (YP10)

Participants suggested that expanding services and extending opening hours could improve wait times and ease of access:

> *“They don’t open on weekends, which I understand is because they only have certain numbers of staff, but, or at night times as well. Because a lot of the time when I struggle, it’s late at night or it’s at the weekend, and so you then have to go to a clinical space.”* (YP5)

Moreover, some young people reported long wait times for therapy, varying from a couple of weeks to six months. Some emphasised the need for greater flexibility in therapy duration, feeling that the limited number of sessions offered (most often between 6-14 sessions maximum) were insufficient for some young people’s needs and led them to access additional support elsewhere:

> *“It would be good if they didn’t have their limit… ‘cause some people like need more sessions than others…that was probably one of the reasons why I was like, oh I’ll go with the NHS.”* (YP4)

An awareness of hubs being over-stretched was also reflected in some young people’s discussions around burdening the service, or staff being overworked:

> *“I think just expanding upon what they have, getting more people, more workers. I know how overworked some of the workers are there and what their caseloads look like, and it’s a lot.”* (YP8).

Some young people felt their hubs should open more locations across the country, particularly in rural or disadvantaged areas. One also felt having more familiar hubs under the same name would help them to access support if they were to relocate:

> *“It would be good… if there were ones dotted around the country, because then, if you know, oh, this, this hub is really good, so therefore, oh, I’m moving up to this place, so I’m going to access that same hub again.”* (YP4)

While some participants felt hub support could be helpful for young people with more complex needs, others felt that it was insufficient:

> *“So the drop in, I used a lot whenever I was feeling like in a crisis. What really helped me is that as well, they didn’t have to define what a crisis is like. I could just use it whatever I felt like I was in a crisis”* (YP5)
>
> *“I would recommend [hub], but I would also recommend looking into transition services in the meantime because obviously 6 to 12 months waiting list is a very long time, especially if you’re in crisis, so I would recommend you know, for more professional and more structured help, absolutely apply, but also have other things in place in the meantime if you are struggling…”* (YP11)

While some participants were satisfied accessing higher-intensity or more specialist support elsewhere, others wanted hubs to offer this. Suggestions for improvements to hubs included employing staff with more specialist expertise and offering more tailored support for neurodivergent individuals (e.g., support groups for those awaiting diagnostic assessments).

A final concern regarding the scale of the hubs related to age thresholds, with participants reaching the upper age limit (generally at 25 years) worrying about the lack of similar open-access holistic services for adults:

> *“In terms of what [hub] does specifically, there’s not that provided for older people and that does make me feel a little nervous.”* (YP2)

## Discussion

In this qualitative study, we interviewed 20 young people aged 16-25 about their experiences of Early Support Hubs in England. Overall, young people endorsed the Early Support Hub model and had mostly positive experiences with the services. They credited the success of the hubs to their easy accessibility, non-clinical and holistic approaches, community feel, and youth-led philosophies. Young people highlighted the need to expand hubs to address high demand and limited capacity, including opening more locations across the country, especially in rural and disadvantaged areas. Furthermore, it was suggested that having more hubs under the same familiar name could improve access. Some participants expressed a desire for hubs to offer longer-term therapy/counselling or more specialist support, while others were satisfied with accessing this from other services alongside hub support. It is possible that such preferences vary, in part, depending on the availability of other services locally and their integration with hubs.

In general, participants felt that hubs improved access to timely wellbeing support, given their low thresholds for entry, straightforward referral processes, and availability of drop-in support. This aligns with priorities outlined in Youth Access‘ ‘Our Minds Our Future Manifesto’ (30).

However, one of the main barriers to access identified by young people in this study was a lack of awareness about hubs and the types of support they offer – partly due to limited promotion, and, in some cases, the buildings’ inconspicuous, residential appearance. Indeed, hubs are not yet widely known nationally, as highlighted by the *Fund the Hubs* campaign, which seeks to increase both awareness and funding (55). Awareness could also be raised through increased promotion and outreach via schools, universities, other wellbeing services, and social media (56), as well as through changes to the physical hubs such as clearer signage, which would make them more noticeable to passersby whilst still maintaining the homely feel that participants valued. Additionally, ensuring hubs are wheelchair and pushchair accessible could help make access more inclusive for all.

Participants viewed their psychological wellbeing as interlinked with broader lifestyle factors, and benefited from the varied emotional, social, and economic support offered at hubs. They stressed the importance of casual conversations with staff about varying difficulties they may be facing at a given visit and liked the non-clinical settings and lack of pressure to pursue formal therapies. This highlights the utility of the ‘one-stop-shop’ approach recommended by NHS England’s Future in Minds Report (31). By providing this varied support, Early Support Hubs appear to aim to both fill gaps left by widespread youth club closures (57) and reduce pressures on other mental health services. While our findings suggest that young people value this dual function of hubs, it may place competing demands on services. International early interventions models vary in how they navigate this. For example, ‘Headspace’ in Australia adopts a holistic approach by integrating early intervention stepped-care mental health support with support for other needs such as employment, education, substance use, and physical health (58). In contrast, ‘Jigsaw’ in Ireland has a narrower remit, focused on providing brief psychological interventions and onward referrals (32). Both models have more defined roles and are coordinated by national organisations that set standards, manage funding, and support consistent delivery across sites, whilst Early Support Hubs in the UK are locally developed and managed, leading to more variation in their design and delivery. The potential benefits of greater national standardisation of Early Support Hubs, such as improved model clarity and consistency, need to be weighed against the value of local flexibility to respond to diverse community needs.

Whilst the flexible and informal approaches of hubs could make them optimally positioned to deliver early intervention for mental health difficulties before symptoms become severe and enduring (6), the extent to which this more generalised model can meet more complex mental health needs is unclear. This is a limitation acknowledged within other support hub models internationally, for example ‘Headspace’ in Australia, where centres in some regions have been enhanced through vertical integration with specialised services for more complex, less common mental health difficulties, including early psychosis (58) and eating disorders (59). Some participants in our study found the drop-in services useful in times of crisis, suggesting hubs provide more than just early intervention or prevention, but others did discuss needing to access more specialised support elsewhere whilst continuing to use hubs on a more casual ad hoc basis. Clarity on what support the hubs can offer, and for what difficulties, may help young people decide the suitability of the services for their needs and help manage their expectations while accessing services.

The youth-centred approach of hubs was also a key contributor to their perceived success. Participants appreciated being in charge of their own support, with their perspectives prioritised over those of parents or other health professionals involved in referrals. Many valued having choice in the extent of their family’s involvement, and a couple expressed a desire for even greater flexibility – for example, by lowering the age threshold for accessing hub counselling without parental consent. Many hubs also regularly consulted youth participation groups for decisions around how their service operates, ensuring young people’s preferences were represented and actioned. These youth-centred approaches may have contributed to the sense of empowerment described by some young people and stood in contrast to participants’ experiences of more pronounced power imbalances in other mental health services – also frequently reported in previous research (60). This youth-led approach at hubs aligns with existing evidence that positioning parents as the primary ‘service user’ can hinder person-centred care (60), that young people want to be heard, have their autonomy respected, and be actively involved in decisions about their care, and that collaborative relationships with clinicians can support their engagement (61). However, as our study only recruited 16–25-year-olds, it is uncertain whether this theme would hold for younger hub users, who may benefit more from family or school involvement in their care. Evidence suggests that parental involvement can enhance treatment outcomes for adolescents (62), and that collaboration between services – such as schools and mental health providers – is generally evaluated positively and seen as helpful and important by professionals, parents/carers, and young people, though evidence for its impact on clinical outcomes is mixed (63,64).

Young people described Early Support Hubs as inclusive spaces, where individuals from diverse backgrounds felt welcome. Some also reported valuing that hub staff had similar backgrounds and shared identities to them. This aligns with previous research suggesting that some, but not all, people prefer therapists with shared identities (e.g., race or ethnicity), and may perceive them more positively (65–67).

Evidence suggests that some marginalised groups continue to experience inequalities in mental healthcare access, experience, and outcomes (68–73). In this study, a participant identified language barriers as a barrier to engaging with hub support, which highlights the need for accommodations such as multilingual support or interpreters for those who are not proficient in English. This point was also raised in the MHPRU’s qualitative interview study with frontline hub staff (in prep), where staff acknowledged the need for such accommodations, including British Sign Language interpreters, but noted a lack of resources to provide them.

Support was described as easily accessible due to simple self-referral processes for therapy and the lack of referrals needed for drop-ins and groups. This may address challenges faced by young people in obtaining external professional referrals, for example due to ineffective GP support (74) or not meeting symptom thresholds for statutory services (75). Flexible scheduling and support delivery methods also enhanced accessibility. However, hubs faced similar challenges of high demand and limited capacity as statutory services, such as CAMHS and primary care mental health services (16), in the NHS and several participants expressed some dissatisfaction with waiting times and the limited number of therapy sessions offered. This is consistent with experiences reported at similar one-stop-shop youth services internationally (76) and suggests a need for more, larger scale, and better-resourced hubs, particularly in rural and disadvantaged areas. These findings highlight the importance of developing a longer-term, standardised plan for scaling up hub provision to ensure consistent access across regions. Nonetheless, hubs did generally provide access to same day drop-in support, even if there was a wait to be seen.

### Strengths and limitations

This is the first qualitative study to explore young people’s experiences and views of Early Support Hubs across multiple sites in England. It captured the experiences of hub users with diverse ages, genders, sexual orientations, and education/employment statuses, including groups that are often under-represented in research. It also benefited from input from young people and a parent with lived experience of mental health services throughout. This helped to enhance the relevance of the findings and ensure that the research reflected their perspectives.

However, this study has some limitations. While we recruited from eight hubs across England, these may not fully reflect the range of hubs nationally. Additionally, we did not systematically compare findings across sites to explore how factors such as hub size, provider types, or local demographics may have influenced the results, which limits our ability to determine the extent to which the findings are context-specific or generalisable. Recruiting directly from hubs with the help of their staff meant young people with more negative experiences may have been less likely to be approached or less willing to participate. Furthermore, several hubs supported recruitment for our study by directly advertising it to their youth advisory groups, which may have resulted in our sample having more positive views about the hub model than other young people who are not as engaged with the services. Most participants had also used their hub for over a year. Purposively recruiting individuals who have used hubs for different lengths of time, or who have lower levels of engagement with hubs, may have facilitated a more in-depth and representative understanding of young people’s experiences and areas for improvement.

Although we purposively sampled young people from minoritised ethnic groups, the sample was predominantly White British (75.0%). Only one participant identified as being from ‘any Asian background’, and four from ‘any Black background’. This may partly reflect the geographical locations of the hubs, many of which were located in areas with predominantly White British populations. There were some other aspects of diversity, such as disability and religion, which we did not collect data on. This might limit our understanding of whether hubs provide appropriate care for groups who may face greater barriers to accessing mental health services.

### Implications for policy, practice, and research

This study highlights young people’s views on both the strengths of Early Support Hubs and areas for improvement, offering important insights for policy, practice, and research. Our findings showed that young people value hubs’ open-access, youth-centred, one-stop-shop approach, as well as their non-clinical environment and ethos, and flexible, holistic, personalised support. Using this model, hubs appear to be able to meet the needs of certain groups of young people, particularly those with less complex mental health needs. Thus, this approach and ethos may be valuable in development of services for young people.

Young people had mixed views on whether hubs should provide more specialist support for those with more complex or higher levels of need, or whether these should be addressed by other services. This underscores the need for greater clarity about the role of hubs within wider mental health service pathways, which may vary by region depending on available resources. The pros and cons of increased standardisation of the early hub support model across locations should be considered, balancing consistency with the need for local flexibility.

Further qualitative research could also explore themes raised by young people in this study in greater depth, for example: approaches to involving families and obtaining parental consent for care, particularly for younger children, those with safeguarding concerns, or children in care; the impact of youth violence and how this can be effectively managed; and how hubs can reach and provide culturally competent care for diverse groups, including marginalised populations such as minoritised ethnic groups, neurodivergent individuals, people with disabilities, and and those with limited English proficiency.

An exploration and comparison of the perspectives of not only young people, but also parents/carers, frontline hub staff, and hub managers is also needed. This could help to develop a theory of change model for hubs, which would clarify how they achieve their intended outcomes, support evaluation by identifying key indicators of success, guide policy and funding decisions by demonstrating impact, and enhance consistency across hubs while allowing for local adaptation. As this study only interviewed young people over the age of 16, it would also be beneficial to explore experiences of hub users who are younger, as well as from a wider range of ethnic backgrounds, in future research to enable a more nuanced exploration of differences in experiences. Understanding this broader range of experiences of young people, families, and staff would be valuable for informing service development.

Quantitative evaluation is also needed to assess whether young people’s perspectives align with measurable outcomes, such as mental health, wellbeing, social and occupational functioning, and satisfaction with support – including over the longer-term. Evaluating cost-effectiveness will be important to inform policymaking and commissioning decisions. Future quantitative research should include control groups (e.g. waiting lists or alternative services) to address limitations of existing studies, which focus on pre-post comparisons (50). Comparing hub users’ demographics with local population data could highlight inequities in access, while exploring any differences in outcomes across groups (e.g., by age, ethnicity, gender identity, sexual orientation, socioeconomic background, geographical location, types of difficulties) could inform strategies to ensure hubs are accessible and responsive to the needs of diverse populations.

## Lived experience commentary by Nima Cas Hunt and Lizzie Mitchell

The reflections in this commentary are derived from our lived experience of using mental health services for young people, including the use of statutory NHS services as well as hub-style, less clinical settings.

This paper highlights the significant improvements in mental health recovery that hubs can generate for young people with ‘common mental health problems’ of anxiety and depression. However, these conditions represent only a small segment of the rise in youth mental health difficulties; whether the needs of young people experiencing more ‘severe’ mental health challenges such as eating disorders, psychosis, OCD, and bipolar disorders are met by the hub is unclear. Although hubs are branded as being accessible, these young people are potentially turned away and left without support, fuelling the stigma that they are complex, othered, and left without the early intervention the hubs promise.

One of the themes in this paper relates to hubs being non-clinical settings which expand on, or depart from, statutory approaches to mental healthcare. Largely, this was perceived positively, with participants tending to view their mental health beyond diagnoses and thus appreciating the non-diagnostically focused, holistic nature of the support. Similarly, many participants emphasised the merit of hubs acting as ‘social spaces’, wherein a consistent, supportive community could be accessed – with further exploration, this theme could potentially challenge what is currently considered best practice in mental healthcare.

To build on this, future research could explore if and how community could be used as an early intervention in tackling mental ill-health in young people, and investigate the effect of the non-clinical support offered by hubs on the young people they serve. Although capturing a range of CYPs’ perspectives, participants in the current paper mainly had positive experiences with the hubs and longer term engagement. Future research would benefit from capturing the experiences of CYPs who have had fewer, or less positive, experiences of the hub, in order to better understand barriers to access, engagement and improve retention rates.

## Data Availability

The data that support the findings of this study are available on request from the corresponding author. The data are not publicly available due to privacy or ethical restrictions.

## Acknowledgements

We would like to acknowledge all the working group members who contributed to this project, including internal MHPRU and external academic, clinical, and lived experience researchers, and give thanks to the participants who gave up their time to take part.

## Appendix A: Topic guide for interviews with service users

*[Start recording]*

### Part 1: Use of early support hubs

1. What sort of difficulties led you to use [hub]?

Potential prompts:

- Would you consider yourself to have any mental health difficulties?

• 2. When did you first and last access a support hub?

• 3. How often have you used [hub]?

Potential prompts:

- Frequency of visits? Eg. once a week, one-off?
- Number of visits overall?

### Part 2: Journey to accessing early support hubs

4. Could you tell me about your journey to receiving support from [hub] for the first time?

Potential prompts:

- How did you find out about [hub]? Eg. recommend by school/friends/family?
- Other support you had accessed/tried to access before [hub]?
- Reasons for choosing [hub] over other services?

5. How easy or difficult was it to access support from [hub]?

Potential prompts:

- Anything that made it easier?
- Anything that made it harder?

### Part 3: Experience of using early support hub

6. Could you tell me about the kind of support you have received from [hub]?

Potential prompts:

- Experiences of drop-in advice/therapy or counselling/groups?
- Did it meet your expectations? Why/why not?

7. How well has the service worked for you?

Potential prompts:

- Parts you particularly liked?
- Parts you didn’t like so much?
- How relevant/sufficient did support feel?
- Any issues you wanted help with that hub didn’t provide support for?
- How well did the location/physical environment work for you?
- What sort of changes in mental health/wellbeing/daily life, if any, did you experience as a result?

8. What were experiences with staff like?

Potential prompts:

- Any issues?

9. To what extent were your family involved in support at [hub]?

Potential prompts:

- Any specific opportunities for family involvement at hub?
- Thoughts/feelings about family being involved/not involved?

10. Reflecting on your overall experiences, would you recommend [hub] to other young people? Why/why not?

### Part 4: Future recommendations for early support hubs

11. How did your experience with [hub] compare to other types of support you have previously used or tried to use?

*(To be asked unless clear from Q4 that there has been no use/attempted use of any other services)*

Potential prompts:

- Any ways hub was more helpful?
- Any ways hub was less helpful?

12. In an ideal world, what would the perfect hub be like?

Potential prompts:

- Process for accessing hub?
- Types of support on offer?
- How support would be delivered?
- What would it physically look like (eg. waiting rooms, support rooms, location)?
- Improvements to current hub that would be needed?

### Part 5: Closing questions

13. Is there anything else you would like to share about your experience of using an early support hub or your views about what works well?

*[End recording]*

